# Trends, patterns and psychological influences on COVID-19 vaccination intention: findings from a large prospective community cohort study in England and Wales (Virus Watch)

**DOI:** 10.1101/2021.03.22.21254130

**Authors:** Thomas Byrne, Parth Patel, Madhumita Shrotri, Sarah Beale, Susan Michie, Jabeer Butt, Nicky Hawkins, Pia Hardelid, Alison Rodger, Anna Aryee, Isobel Braithwaite, Wing Lam Erica Fong, Ellen Fragaszy, Cyril Geismar, Jana Kovar, Annalan M D Navaratnam, Vincent Nguyen, Andrew Hayward, Robert W Aldridge, Virus Watch Collaborative.

**Affiliations:** Centre for Public Health Data Science, Institute of Health Informatics, University College London, UK; Institute of Epidemiology and Health Care, University College London, London, UK; Centre for Behaviour Change, University College London, London, UK; Race Equality Foundation, UK; Independent Consultant, UK; Department of Population, Policy and Practice, UCL Great Ormond Street Institute of Child Health, London, UK; Institute for Global Health, University College London, London, UK; Royal Free London NHS Foundation Trust, London, UK; Department of Infectious Disease Epidemiology, London School of Hygiene and Tropical Medicine, Keppel Street, London, UK; Francis Crick Institute, London, UK; Health Protection and Influenza Research Group, Division of Epidemiology and Public Health, University of Nottingham School of Medicine, Nottingham, United Kingdom; University College London Hospital, London, United Kingdom; Department of Computer Science, University College London, London, UK; London Centre for Nanotechnology and Division of Medicine, London, UCL; SpaceTimeLab, Department of Civil, Environmental and Geomatic Engineering, University College London, London, UK

## Abstract

**Objectives:** To assess trends in intention to accept a COVID-19 vaccine between 1 December 2020 and 25 February 2021, explore associations between socio-demographic factors and vaccination intention and investigate how COVID-19 vaccine- and illness-related attitudes, beliefs and emotions influence vaccination intention.

**Design:** Prospective household community cohort study of COVID-19 infection (Virus Watch).

**Settin:** Online survey of Virus Watch study participants in the community across England and Wales.

**Participants:** Individuals could enrol in Virus Watch if all household members agreed to participate and at least one household member had access to the internet, an email address, and could read English. All Virus Watch participants aged 16 years and over who responded to questions relating to COVID-19 vaccine intention in questionnaires between December 2020 and February 2021 were included in this analysis.

**Main outcome measures:** Vaccination intention was measured by individual participant responses to ‘Would you accept a COVID-19 vaccine if offered?’, collected between 1-14 December 2020 and 17-25 February 2021. Possible responses were ‘Yes’, ‘No’ and ‘Unsure’ (December 2020 &February 2021) and ‘Already had a COVID-19 vaccine’ (February 2021 only). Responses to a 13-item questionnaire collected between 4-11 January 2021 were analysed using factor analysis to investigate psychological influences (attitudes, beliefs and emotions) on vaccination intention.

**Results:** Survey response rate was 56% (20,792/36,998) in December 2020 and 52% (20,284/38,727) in February 2021, with 14,713 adults reporting across both time points. Of participants reporting across both timepoints, 13,281 (90%) answered ‘Yes’ and 1,432 (10%) responded ‘No’ or ‘Unsure’ in December 2020. Of those answering ‘No’ or ‘Unsure’ in December 2020, 1,233 (86%) went on to answer ‘Yes’ or ‘Already had a COVID-19 vaccine’ in February 2021. The magnitude of this shift was consistent across all ethnic groups measured and all levels of social deprivation. Age was most strongly associated with vaccination intention, with 16–24-year-olds more likely to respond “No” or “Unsure” than those aged 75+ in December 2020 (RR: 4.32, 95% CI: 2.40-7.78 &2.93 95% CI: 2.19-3.92, respectively) and February 2021 (RR: 5.30 95% CI: 1.39-20.20 &20.21 95%CI: 7.19-56.78). The association between ethnicity and vaccination intention has weakened, but not disappeared, over time. Both vaccine- and illness-related psychological factors were shown to influence vaccination intention.

**Conclusions:** Over four in five adults (86%) who were reluctant or intending to refuse a COVID-19 vaccine in December 2020 had changed their mind in February 2021 and planned on accepting, or had already accepted, a vaccine.

## Introduction

Alongside availability and delivery, vaccination intention, which refers to the intention to take or refuse a vaccine when offered, determines the success of a vaccination programme. In 2019, the World Health Organization listed the reluctance or refusal of vaccines as one of the top threats to global public health.^1^ Patterns and drivers of vaccination intention vary over time and region, correlating to local politics, history and religion.^2^

The UK’s COVID-19 vaccination programme plans to achieve high levels of vaccination coverage across the population.^3^ Diverse supply chains and delivery through primary care services has meant the UK currently has one of the highest COVID-19 vaccination rates in the world.^4^ Public intention to take a COVID-19 vaccine when offered is high in the UK,^5^ but early evidence of disparities between ethnic and social groups has led to significant concern among public health practitioners, the NHS (which is leading the UK’s vaccine delivery), voluntary sector organisations, and politicians.^6–8^

Previous studies have found that age, ethnicity, income and education are independently associated with COVID-19 vaccination intention.^9,10^ More specifically, they find that young adults, people from most minority ethnic backgrounds, people on low income and people with low education levels are more likely to be reluctant or refuse a COVID-19 vaccine. It is likely that reluctance or refusal to take a vaccine is contributing to the disparities in COVID-19 vaccination coverage, which is lower in most minority ethnic populations and in areas of higher deprivation.^11,12^ However, it is unclear to what extent disparities in COVID-19 vaccination coverage are a result of differences vaccination intention as opposed to structural factors that determine access to healthcare services.^13,14^

Evidence gathered before the UK COVID-19 vaccination programme commenced suggested that beliefs around the efficacy and safety of COVID-19 vaccines are likely to be the greatest psychological influence on vaccination intention.^15^ It is not clear if this remained true since the COVID-19 vaccination programme began and if beliefs and perceptions around the risk of COVID-19 illness also influence vaccination intention.

Disparities in COVID-19 vaccination coverage are especially concerning given the greater risk of COVID-19 infection, severe illness and death in most minority ethnic populations and areas of high deprivation.^16,17^ Following concerted and targeted action to increase public intention to take a COVID-19 vaccine, it is not yet clear how, or whether, vaccination intention is changing over time in England and Wales.

This study aims to:

1. Examine whether and how COVID-19 vaccination intention has changed over time, across different populations in England and Wales
2. Investigate socio-demographic factors associated with current vaccination intention in England and Wales
3. Investigate how vaccine- and illness-related psychological factors (attitudes, beliefs and emotions) may influence vaccination intention and whether these factors vary across populations

## Methods

### Study design and procedure

Data for this analysis was collected as part of the Virus Watch study, a large prospective household community cohort study of the transmission and burden of COVID-19 in England and Wales. The full study design and methodology has been described elsewhere.^18^ Data collection using online REDCap surveys began on 24 June 2020 and is ongoing.

After enrolling in the study, an initial baseline survey collected demographic, occupation, financial and medical history data from participants. Thereafter, participants were surveyed weekly (contacted by email) on the presence or absence of symptoms that could indicate COVID-19 disease, activities undertaken prior to symptom onset, any SARS-CoV-2 swab test results, and COVID-19 vaccine uptake in the previous week. Bespoke monthly surveys collected detailed information on potential determinants of SARS-CoV-2 infection and COVID-19 disease.

### Participants

Participants were recruited into the Virus Watch study using a range of methods including by post, social media, SMS messages or personalised letters with incentives from General Practices. Participants were eligible if all household members agreed to take part, if they had access to the internet (Wi-Fi, fixed or on a mobile phone) and an email address. At least one household member had to be able to read English to complete the surveys. Participants were not eligible if their household was larger than 6 people (due to limitations of the online survey infrastructure).

In the analyses described in this report, we excluded respondents under the age of 16 for two reasons. First, survey responses are more likely to represent parental views than views of children themselves as the surveys may be completed by an adult on their child’s behalf. Second, children are not yet eligible for vaccination in the UK, which may have influenced parents’ intention to vaccinate their children.

### Exposures

We explored key demographic, social, and clinical variables that could be associated with COVID-19 vaccination intention amongst adults. Age (on entry to the study) and sex (at birth) were defined *a priori* as variables of interest. Other variables of interest included self-reported ethnicity, grouped as per the following ONS categories: ‘White British’, ‘White Irish’, ‘White Other’, ‘South Asian’, ‘Other Asian’, ‘Black’, ‘Mixed’, ‘Other ethnicity’; whether born in the UK or born abroad; region of residence within England and Wales; small area-level deprivation using the Indices of Multiple Deprivation (IMD) based on postcode of residence;^19^ presence of comorbidities associated with higher risk of adverse COVID-19 outcomes (as defined by Public Health England - see Appendix IA) based on data from the baseline survey; health or care worker status; and inclusion within one of the UK Joint Committee on Vaccination and Immunisation (JCVI)’s priority groups for COVID-19 vaccination (see Appendix IA).^20^ Data on exposure variables were collected through the baseline survey completed on entry into the Virus Watch study.

### Outcomes

The primary outcome of interest was each participant’s response to the following question: *‘Would you accept a COVID-19 vaccine if offered?’*. Outcome data were collected over two separate time periods (1-14 December 2020 and 17-25 Feb 2021) through surveys sent to the whole Virus Watch cohort on the first listed day. Possible responses were: *‘Yes’, ‘No’, ‘Unsure’* (December and February surveys), and *‘Already had a COVID-19 vaccine’* (February survey only).

Between 4th-11th January 2021, psychological influences on vaccination intention were surveyed using a 13-item questionnaire measuring attitudes, beliefs and emotions related to COVID-19 illness and vaccination. This questionnaire was adapted from a 16-item measure used in the Flu Watch cohort study to measure vaccination-related attitudes during the H1N1 influenza pandemic;^15^ three items were removed, and the wording of the remaining items adapted to reflect the current pandemic situation. Participants rated their agreement for all items on a 5-point Likert scale (‘strongly disagree’ – ‘strongly agree’), with negative-worded items reverse coded prior to analysis. The full questionnaire is provided in Appendix II.

### Statistical analysis

Baseline and monthly survey response data were extracted from REDCap, linked, and analysed in Stata (version 16.0, StataCorp). Observations missing data on the primary outcome of interest were excluded from the denominators of all analyses. Respondents with missing baseline information were excluded from only the relevant explanatory variable denominator; for example, those missing a postcode were excluded from analyses by region and IMD.

To examine changes in intention over time, responses to the February survey were compared to those from December among individuals with complete data at both time points. We grouped ‘No’ and ‘Unsure’ responses due to small numbers. The percentage change in response was calculated overall and by ethnicity and IMD quintile. We suppressed low cell counts across certain categories to reduce the possibility of deductive disclosure.

We used mixed-effects Poisson regression with robust standard errors to derive risk ratios and 95% confidence intervals for the association between a range of demographic, social, and clinical participant characteristics and responses to the question ‘Would you accept a COVID-19 vaccine if offered?’ in the December 2020 and February 2021 surveys. In the first model we compared those responding ‘No’ with those responding ‘Yes’, and in the second model we compared those responding ‘Unsure’ with those responding ‘Yes’. Guided by the Virus Watch study’s community advisory group, we investigated these responses separately to illustrate any differences in factors associated with uncertainty versus those associated with intention to refuse. We first tested the association for each variable separately, adjusting only for age and sex (*a priori* variables). Variables that were significantly associated in univariable analyses and considered theoretically relevant to vaccine intention were included in the fully-adjusted multivariable models of both outcomes (“Yes” vs “No” and “Yes” vs “Unsure”). The multivariable models also included a random term to account for clustering at the household level.

To identify psychological influences on vaccination intention, survey responses from the January monthly survey were split into a ‘training’ dataset (*n*=10088 responses) for exploratory factor analysis and a ‘cross-validation’ dataset (*n=*10890) for confirmatory factor analysis.

Further details of the approach are in Appendix IV. To assess whether psychological factors differed by socio-demographic characteristics, we compared median factor scores based on their exact 95% confidence intervals by age group, sex, IMD quintile, and ethnicity.

### Patient and public involvement

The study team worked with the Race Equality Foundation and Doctors of the World who advised on the inclusion of people from minority ethnic backgrounds in Virus Watch and set up a community advisory group to inform the ongoing design and dissemination of health equity aspects of Virus Watch. This advisory group, consisting of lay members of the public, community leaders, charities and policy organisation, guided and reviewed the analyses described in this paper.

## Results

When data were extracted on 25 February 2021, there were 22,556 households and 46,539 people taking part in Virus Watch across England and Wales, of whom 40,797 were adults aged 16 years and over and 4,412 (11%) people from minority ethnic backgrounds (Table 1).

**Table 1.**
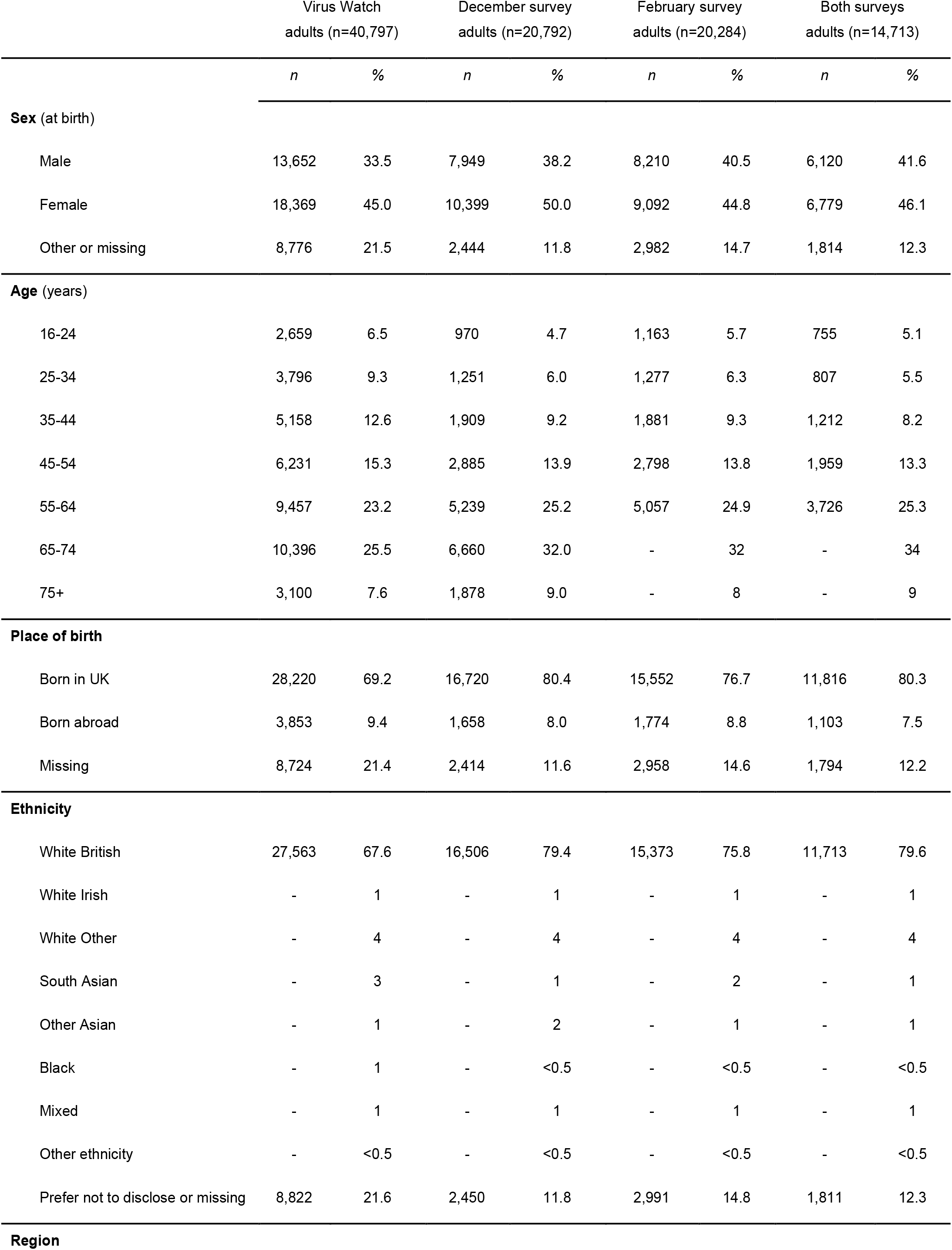
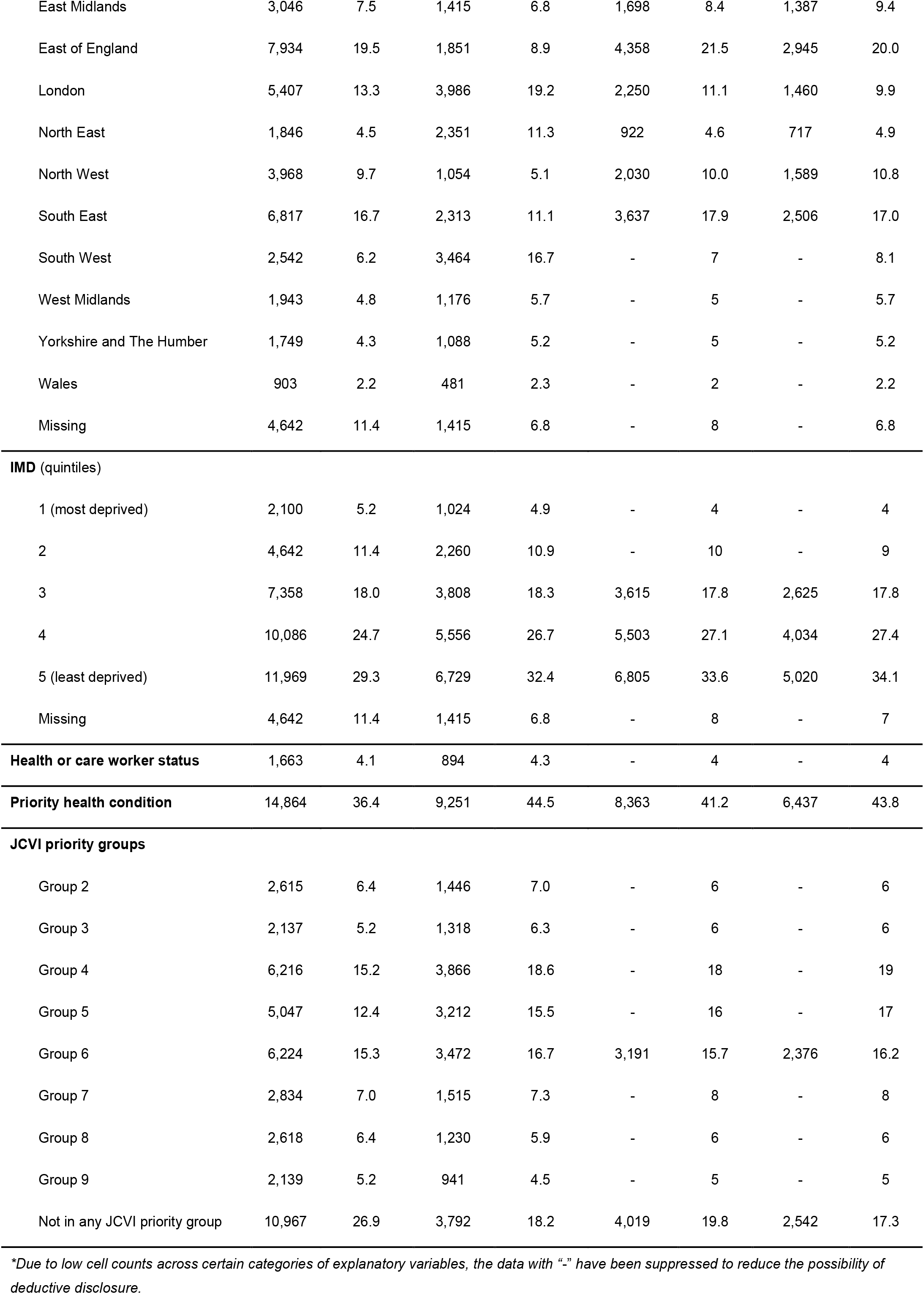
Characteristics of adult Virus Watch study participants (25th February 2021) and those responding to the question on COVID-19 vaccine acceptance in December 2020, in February 2021, and in both surveys.

|  | Virus Watch<br>adults (n=40,797) |  | December survey<br>adults (n=20,792) |  | February survey<br>adults (n=20,284) |  | Both surveys<br>adults (n=14,713) |  |
| --- | --- | --- | --- | --- | --- | --- | --- | --- |
|  | *n* | % | *n* | % | *n* | % | *n* | % |
| **Sex (at birth)** |  |  |  |  |  |  |  |  |
| Male | 13,652 | 33.5 | 7,949 | 38.2 | 8,210 | 40.5 | 6,120 | 41.6 |
| Female | 18,369 | 45.0 | 10,399 | 50.0 | 9,092 | 44.8 | 6,779 | 46.1 |
| Other or missing | 8,776 | 21.5 | 2,444 | 11.8 | 2,982 | 14.7 | 1,814 | 12.3 |
| **Age (years)** |  |  |  |  |  |  |  |  |
| 16-24 | 2,659 | 6.5 | 970 | 4.7 | 1,163 | 5.7 | 755 | 5.1 |
| 25-34 | 3,796 | 9.3 | 1,251 | 6.0 | 1,277 | 6.3 | 807 | 5.5 |
| 35-44 | 5,158 | 12.6 | 1,909 | 9.2 | 1,881 | 9.3 | 1,212 | 8.2 |
| 45-54 | 6,231 | 15.3 | 2,885 | 13.9 | 2,798 | 13.8 | 1,959 | 13.3 |
| 55-64 | 9,457 | 23.2 | 5,239 | 25.2 | 5,057 | 24.9 | 3,726 | 25.3 |
| 65-74 | 10,396 | 25.5 | 6,660 | 32.0 | - | 32 | - | 34 |
| 75+ | 3,100 | 7.6 | 1,878 | 9.0 | - | 8 | - | 9 |
| **Place of birth** |  |  |  |  |  |  |  |  |
| Born in UK | 28,220 | 69.2 | 16,720 | 80.4 | 15,552 | 76.7 | 11,816 | 80.3 |
| Born abroad | 3,853 | 9.4 | 1,658 | 8.0 | 1,774 | 8.8 | 1,103 | 7.5 |
| Missing | 8,724 | 21.4 | 2,414 | 11.6 | 2,958 | 14.6 | 1,794 | 12.2 |
| **Ethnicity** |  |  |  |  |  |  |  |  |
| White British | 27,563 | 67.6 | 16,506 | 79.4 | 15,373 | 75.8 | 11,713 | 79.6 |
| White Irish | - | 1 | - | 1 | - | 1 | - | 1 |
| White Other | - | 4 | - | 4 | - | 4 | - | 4 |
| South Asian | - | 3 | - | 1 | - | 2 | - | 1 |
| Other Asian | - | 1 | - | 2 | - | 1 | - | 1 |
| Black | - | 1 | - | <0.5 | - | <0.5 | - | <0.5 |
| Mixed | - | 1 | - | 1 | - | 1 | - | 1 |
| Other ethnicity | - | <0.5 | - | <0.5 | - | <0.5 | - | <0.5 |
| Prefer not to disclose or missing | 8,822 | 21.6 | 2,450 | 11.8 | 2,991 | 14.8 | 1,811 | 12.3 |
| **Region** |  |  |  |  |  |  |  |  |
| East Midlands | 3,046 | 7.5 | 1,415 | 6.8 | 1,698 | 8.4 | 1,387 | 9.4 |
| East of England | 7,934 | 19.5 | 1,851 | 8.9 | 4,358 | 21.5 | 2,945 | 20.0 |
| London | 5,407 | 13.3 | 3,986 | 19.2 | 2,250 | 11.1 | 1,460 | 9.9 |
| North East | 1,846 | 4.5 | 2,351 | 11.3 | 922 | 4.6 | 717 | 4.9 |
| North West | 3,968 | 9.7 | 1,054 | 5.1 | 2,030 | 10.0 | 1,589 | 10.8 |
| South East | 6,817 | 16.7 | 2,313 | 11.1 | 3,637 | 17.9 | 2,506 | 17.0 |
| South West | 2,542 | 6.2 | 3,464 | 16.7 | - | 7 | - | 8.1 |
| West Midlands | 1,943 | 4.8 | 1,176 | 5.7 | - | 5 | - | 5.7 |
| Yorkshire and The Humber | 1,749 | 4.3 | 1,088 | 5.2 | - | 5 | - | 5.2 |
| Wales | 903 | 2.2 | 481 | 2.3 | - | 2 | - | 2.2 |
| Missing | 4,642 | 11.4 | 1,415 | 6.8 | - | 8 | - | 6.8 |
| IMD (quintiles) |  |  |  |  |  |  |  |  |
| 1 (most deprived) | 2,100 | 5.2 | 1,024 | 4.9 | - | 4 | - | 4 |
| 2 | 4,642 | 11.4 | 2,260 | 10.9 | - | 10 | - | 9 |
| 3 | 7,358 | 18.0 | 3,808 | 18.3 | 3,615 | 17.8 | 2,625 | 17.8 |
| 4 | 10,086 | 24.7 | 5,556 | 26.7 | 5,503 | 27.1 | 4,034 | 27.4 |
| 5 (least deprived) | 11,969 | 29.3 | 6,729 | 32.4 | 6,805 | 33.6 | 5,020 | 34.1 |
| Missing | 4,642 | 11.4 | 1,415 | 6.8 | - | 8 | - | 7 |
| Health or care worker status |  |  |  |  |  |  |  |  |
|  | 1,663 | 4.1 | 894 | 4.3 | - | 4 | - | 4 |
| Priority health condition |  |  |  |  |  |  |  |  |
|  | 14,864 | 36.4 | 9,251 | 44.5 | 8,363 | 41.2 | 6,437 | 43.8 |
| JCVI priority groups |  |  |  |  |  |  |  |  |
| Group 2 | 2,615 | 6.4 | 1,446 | 7.0 | - | 6 | - | 6 |
| Group 3 | 2,137 | 5.2 | 1,318 | 6.3 | - | 6 | - | 6 |
| Group 4 | 6,216 | 15.2 | 3,866 | 18.6 | - | 18 | - | 19 |
| Group 5 | 5,047 | 12.4 | 3,212 | 15.5 | - | 16 | - | 17 |
| Group 6 | 6,224 | 15.3 | 3,472 | 16.7 | 3,191 | 15.7 | 2,376 | 16.2 |
| Group 7 | 2,834 | 7.0 | 1,515 | 7.3 | - | 8 | - | 8 |
| Group 8 | 2,618 | 6.4 | 1,230 | 5.9 | - | 6 | - | 6 |
| Group 9 | 2,139 | 5.2 | 941 | 4.5 | - | 5 | - | 5 |
| Not in any JCVI priority group | 10,967 | 26.9 | 3,792 | 18.2 | 4,019 | 19.8 | 2,542 | 17.3 |

*\*Due to low cell counts across certain categories of explanatory variables, the data with “-” have been suppressed to reduce the possibility of deductive disclosure.*

### 1. Trends in vaccination intention

Participants responded to the survey question “Would you accept a COVID-19 vaccine if offered?”. The response rate for participants aged 16 or over to the bespoke monthly survey in December 2020 was 56% (20,792/36,998) and in February 2021 was 52% (20,284/38,727), with 14,713 adults reporting across both time points. Table 1 summarises the characteristics of people who answered these surveys.

In December 2020, 18,517 (89%) participants responded “Yes”; 1,813 (9%) said they were “Unsure”; and 462 (2%) responded “No”. In February 2021, 7,778 (38%) participants responded “Already had a COVID-19 vaccine”; 12,039 (59%) responded “Yes”; 284 (1%) said they were “Unsure”; and 183 (1%) responded “No” (see Appendix IB for full description of responses by explanatory variables).

Examining only participants who answered the survey question at both timepoints, 13,281 responded “Yes” in December 2020. Of these, 13,190 (99%) went on to respond “Yes” or “Already had a COVID-19 vaccine” and 91 (1%) to respond “No” or “Unsure” in February 2021 (Table 2).

**Table 2:** Change in responses to “Would you accept a COVID-19 vaccine if offered” from December to February, by ethnicity and IMD quintile*

|  | "Yes" in Dec 2020<br>N=13,281 (90%) when asked again in Feb 2021 |  | "No or Unsure" in Dec 2020<br>N=1,432 (10%) when asked again in Feb 2021 |  |
| --- | --- | --- | --- | --- |
|  | Yes or Already had<br>N=13,190 (99%) | No or Unsure<br>N=91 (1%) | Yes or Already had<br>N=1,233 (86%) | No or Unsure<br>N=199 (14%) |
| **Ethnicity category (n=12,902)** |  |  |  |  |
| White British | 10,650 (99%) | 61 (1%) | 876 (87%) | 126 (13%) |
| White Irish | - (98%) | - (2%) | - (78%) | - (22%) |
| White Other | - (99%) | - (1%) | - (78%) | - (22%) |
| South Asian | - (98%) | - (2%) | - (90%) | - (10%) |
| Other Asian | - (99%) | - (1%) | - (73%) | - (27%) |
| Black | - (96%) | - (4%) | - (88%) | - (13%) |
| Mixed | - (98%) | - (2%) | - (72%) | - (28%) |
| Other Ethnicity | - (100%) | - (0%) | - (79%) | - (21%) |
| **IMD Quintile (n=13,715)** |  |  |  |  |
| 1 (Most deprived) | - (99%) | - (1%) | - (83%) | - (17%) |
| 2 | - (99%) | - (1%) | - (79%) | - (21%) |
| 3 | 2,338 (99%) | 21 (1%) | 229 (86%) | 37 (14%) |
| 4 | 3,656 (99%) | 22 (1%) | 307 (86%) | 49 (14%) |
| 5 (Least deprived) | 4,597 (99%) | 24 (1%) | 357 (89%) | 42 (11%) |

\*Due to low cell counts across certain categories of ethnicity and IMD, the data with "-" have been suppressed to reduce the possibility of deductive disclosure.

Again, examining only participants who answered the survey question at both timepoints, 1,432 responded “No” or “Unsure” in December 2020. Of these, 1,233 (86%) went on to respond “Yes” or “Already had a COVID-19 vaccine” in February 2021. 199 (14%) responded “No” or “Unsure” once again in February 2021. This shift in the intention to accept a COVID-19 vaccine was observed at a similar magnitude across all ethnic groups measured ranging from 72% in people from Mixed ethnic backgrounds to 90% in people from South Asian ethnic backgrounds (Table 2). It is also consistent across all IMD quintiles ranging from 79% to 89%. Where cell sizes are less than 10, percentages are presented without denominators to minimise the possibility of deductive disclosure.

### 2. Factors associated with uncertainty or intention to refuse vaccination

Sex at birth, age group, ethnicity, IMD quintile and having a priority health condition were included in all fully-adjusted multivariable regression models (Figure 1). In the final models, age was most strongly associated with vaccination intention in both models, with low intention to accept a COVID-19 vaccine inversely related to age. In the February survey 25–35-year-olds were over eight times (RR 8.58, 95%CI 2.35, 31.4) more likely to say ‘No’ and 16-24-year-olds were twenty times (RR 20.02, 95%CI 7.19, 56.8) more like to say ‘Unsure’ compared with over-75-year-olds.

**Figure 1A:**
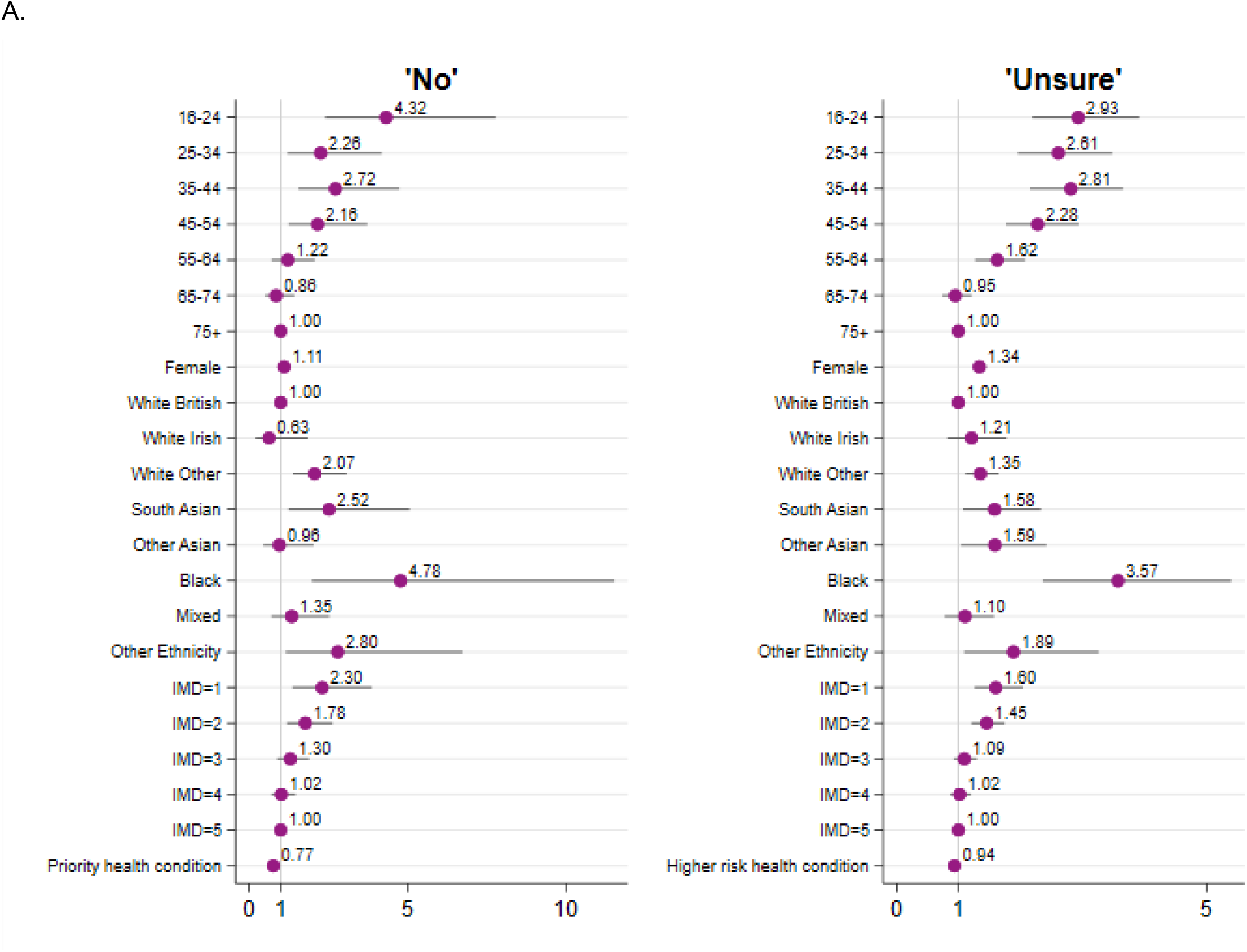
Risk ratio estimates and 95% confidence intervals from multivariable regression models exploring factors associated with intention to accept a COVID-19 vaccine (‘No’ or ‘Unsure’ compared with ‘Yes’) in December 2020 (see tables in Appendix IB, IC). Estimates are adjusted for all other variables in the model.

**Figure 1B:**
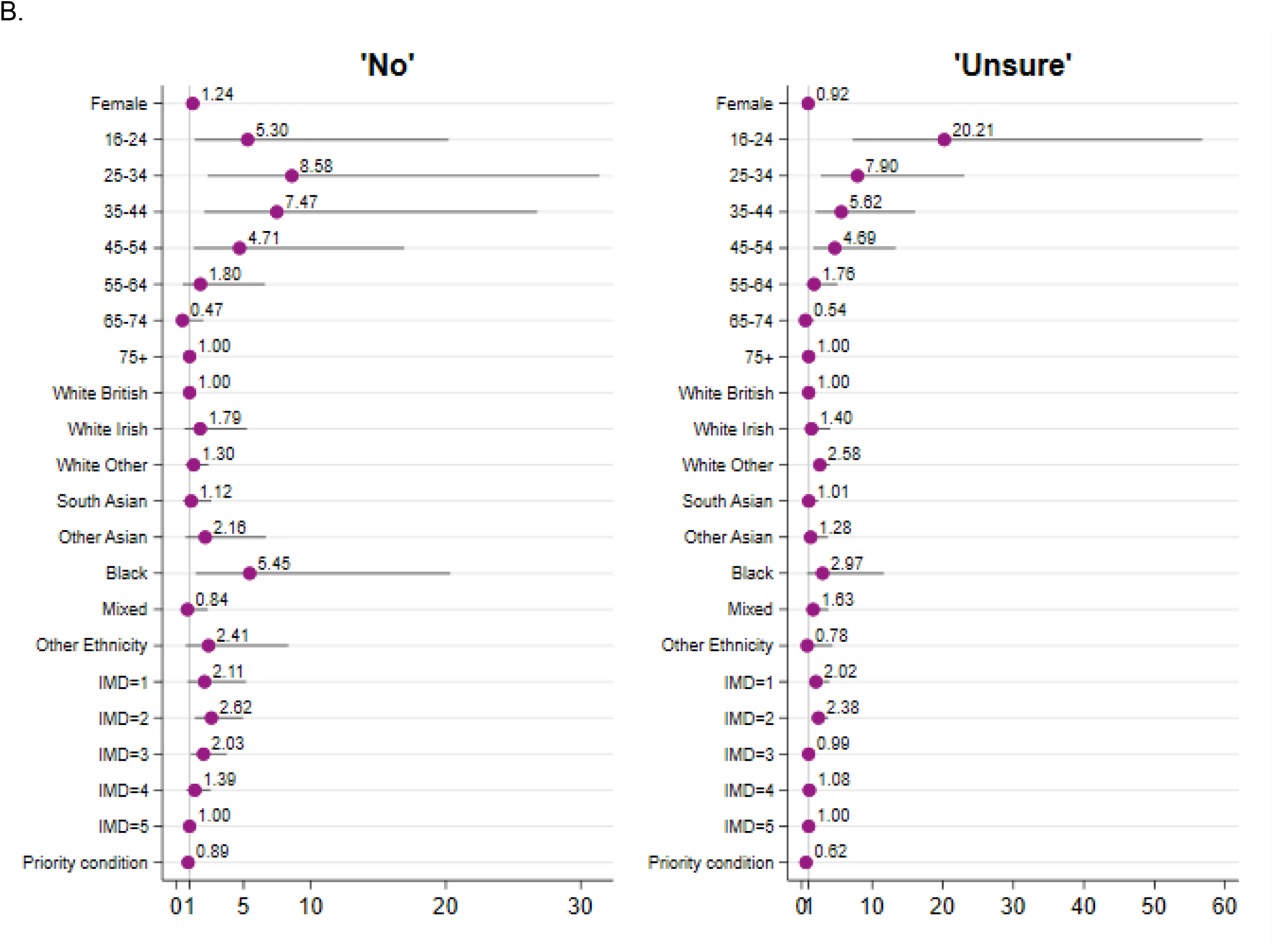
Risk ratio estimates and 95% confidence intervals from multivariable regression models exploring factors associated with intention to accept a COVID-19 vaccine (‘No’ or ‘Unsure’ compared with ‘Yes’) in February 2021 (see tables in Appendix IB, IC). Estimates are adjusted for all other variables in the model.

In December 2020, females were more likely to respond ‘Unsure’ compared to males (RR 1.34, 95% CI 1.24, 1.44). There was no significant difference by sex observed in February 2021. People with a health condition that is prioritised for COVID-19 vaccination (as defined by Public Health England - see Appendix IA) are more likely to respond ‘Yes’ across both time points.

In December 2020, ethnicity was associated with vaccine intention after adjustment for all other explanatory variables. People from White Other (‘No’: RR 2.07, 95%CI 1.38, 3.08, p<0.001; ‘Unsure’: RR 1.35, 95%CI 1.11, 1.65, p<0.001), South Asian (‘No’: RR 2.52, 95%CI 1.26, 5,06, p=0.009; ‘Unsure’: RR 1.58, 95%CI 1.07, 2.33, p=0.021), Black (‘No’: RR 4.78, 95%CI 1.98, 11.51, p<0.001; ‘Unsure’: RR 3.57, 95%CI 2.36, 5.40, p<0.001), and Other (‘No’: RR 2.8, 95%CI 1.16, 6.74, p=0.022; ‘Unsure’: RR 1.89, 95%CI 1.09, 3.26, p=0.023) ethnic backgrounds more likely to say ‘No’ and ‘Unsure’ compared with those from White British backgrounds. In February 2021, this association was much weaker and disparities in vaccine intention between ethnic groups were reduced. Only people from Black ethnic backgrounds (RR 5.45, 95%CI 1.46, 20.3, p=0.012) remained more likely to say ‘No’ compared to White British people, and only people from White Other ethnic backgrounds (RR 2.58, 95%CI 1.67, 3.98) remained more likely to say ‘Unsure’ compared to White British people.

Across both time points, there was a gradient in vaccine intention by local area deprivation. In December 2020, people living in the two most deprived quintiles (IMD=1 and IMD=2) were more likely to say ‘No’ and ‘Unsure’ compared to those living in the least deprived quintile (IMD=5). In February 2021, only those from IMD=2 were more likely to say ‘No’ and ‘Unsure’ (‘No’: RR 2.62, 95%CI 1.38, 4.97, p=0.003; ‘Unsure’: RR 2.38, 95%CI 1.5, 3.77, p<0.001) compared to those living in least deprived quintile.

### 3. Psychological influences on vaccination intention

Exploratory factor analysis (EFA) of responses to the 13-item questionnaire suggested a two-factor structure. Factor 1 comprises beliefs and concerns around the efficacy, safety, side-effects, and time burden of vaccination and is made up of eight questions (items 6-13 in questionnaire, see supplementary materials), e.g. “I am concerned that the COVID-19 vaccine will not have been tested enough”. Factor 2 comprises risk perception and concern around acquiring, transmitting, and suffering severe effects of COVID-19 and is made up of four questions (items 1-4 in questionnaire), e.g. “I do not think that I am at risk of COVID-19”. A single item (item 5: “worry about time off work/education due to COVID-19”) was removed during EFA due to improved scale reliability after removal (Cronbach’s α 0.75 vs 0.78), and low communality and factor loadings. This item was considered theoretically relevant and retained as an individual item in later analyses. Removal of this item did not substantially affect the EFA results. All indices of relative and absolute fit based on confirmatory factor analysis (CFA) indicated that the two-factor EFA model was the best fit to the data, compared to univariable models including or excluding Item 5. Detailed EFA results pre- and post-removal and CFA results including factor loadings and indices of relative and absolute fit are reported in Appendix III.

Factors 1 and 2 were found to consistently predict responding “Unsure” or “No” when participants were asked about their vaccination intention (Table 3). The first factor, relating to COVID-19 vaccines, demonstrated the strongest association with being unsure about vaccination (December RR: 0.29 [0.27, 0.31]; February RR: 0.20 [0.17, 0.24]) and intended refusal (December RR: 0.22 [0.20, 0.25]; February RR: 0.25 [0.20, 0.31]). The second factor, relating to COVID-19 illness, was also predictive of being unsure (December RR: 0.64 [0.60, 0.68]; February RR: 0.35 [0.29, 0.41]) and intended refusal (December RR: 0.41 [0.36, 0.47]; February RR: 0.43 [0.32, 0.57]). Worries about missing work or education due to COVID-19 (item 5), for which analyses were limited to participants who reported being in work or education, were associated with an increased risk of being unsure about taking a COVID-19 vaccine (RR:1.06 [1.01, 1.12]), but not intended refusal (RR: 1.04 [0.92, 1.17]) in December only.

**Table 3.** Risk Ratios between Psychological Factors and COVID-19 Vaccine Intention

|  | Risk Ratio (95% CI) |  |  |  |
| --- | --- | --- | --- | --- |
|  | December |  | February |  |
|  | Unsure | No | Unsure | No |
| Factor 1 - Beliefs and concerns around vaccination | 0.29<br>(0.27, 0.31) | 0.22<br>(0.20, 0.25) | 0.20<br>(0.17, 0.24) | 0.25<br>(0.20, 0.31) |
| Factor 2 - Illness-related risk perception and concern | 0.64<br>(0.60, 0.68) | 0.41<br>(0.36, 0.47) | 0.35<br>(0.29, 0.41) | 0.43<br>(0.32, 0.57) |
| Item 5 - 'I was worried about having to take time off work/education because of COVID-19' | 1.06<br>(1.01, 1.12) | 1.04<br>(0.92, 1.17) | 1.16<br>(0.97, 1.40) | 1.16<br>(0.95, 1.43) |

\*limited to participants in work/education at baseline

Both psychological factors varied by age (Table 4), with more positive vaccination-related attitudes and greater concern about COVID-19 illness in older age groups, based on Factors 1 and 2 (e.g., for both factors, median for 16-24 years: 4, 95% CI: 4,4; 75+ years: 5 95% CI: 5, 5). White British participants had higher median scores (5, 95% CI: 5, 5) for Factor 1, indicating more positive views about COVID-19 vaccines compared to participants from several minority ethnic minority groups, including participants identifying as Black (4, 95%CI: 3.5, 4), South Asian (4, 95%CI: 4, 4.5), Other Asian (4, 95%CI: 4, 4.5), White Other (4.5, 95%CI: 4.5, 4.5), and Other Ethnicity (4, 95%CI: 4, 4.93). Perceptions and concerns about COVID-19 illness did not differ by ethnicity.

**Table 4.** Median Factor Scores by Socio-Demographic Characteristics

|  |  | Median (95% CI) |  |  |
| --- | --- | --- | --- | --- |
|  |  | Factor 1 | Factor 2 | Item 5 |
|  |  | Beliefs and concerns around vaccination | Illness-related risk perception and concern | 'I was worried about having to take time off work/education because of COVID-19'* |
| Age Group | 16-24 | 4 (4-4) | 4 (4, 4) | 4 (3, 4) |
|  | 25-34 | 4.5 (4.5-4.5) | 4 (4, 4) | 3 (3, 3) |
|  | 35-44 | 4.5 (4, 4.5) | 4 (4, 4) | 3 (3, 3) |
|  | 45-54 | 4.5 (4.5, 4.5) | 4.5 (4.5, 4.5) | 3 (3, 3) |
|  | 55-64 | 5 (5, 5) | 4.5 (4.5, 4.5) | 3 (3, 3) |
|  | 65-74 | 5 (5,5) | 5 (5, 5) | 2 (2, 3) |
|  | 75+ | 5 (5, 5) | 5 (5, 5) | 2 (2, 3) |
| Sex | Male | 5 (4.5, 5) | 4.5 (4.5, 4.5) | 3 (3, 3) |
|  | Female | 5 (5, 5) | 4.5 (4.5, 4.5) | 3 (3, 3) |
| Ethnicity | White British | 5 (5, 5) | 4.5 (4.5, 4.5) | 3 (3, 3) |
|  | White Irish | 5 (4.5, 5) | 4.5 (4.5, 5) | 3 (2, 4) |
|  | White Other | 4.5 (4.5, 4.5) | 4.5 (4.5, 4.5) | 3 (3, 3) |
|  | South Asian | 4 (4, 4.5) | 4.5 (4.5, 5) | 3 (3, 4) |
|  | Other Asian | 4 (4, 4.5) | 4.5 (4, 4.5) | 3 (3, 4) |
|  | Black | 4 (3.5, 4) | 4.5 (4, 5) | 2 (1.33,3) |
|  | Mixed | 4.5 (4, 5) | 4.5 (4, 4.5) | 3 (2, 4) |
|  | Other Ethnicity | 4 (4, 4.93) | 5 (4, 5) | 3 (2.13,4) |
| IMD | 1 (Most deprived) | 4.5 (4.5, 5) | 4.5 (4.5, 4.5) | 3 (3, 3) |
|  | 2 | 4.5 (4.5, 5) | 4.5 (4.5, 4.5) | 3 (3, 3) |
|  | 3 | 5 (4.5, 5) | 4.5 (4.5, 4.5) | 3 (3, 3) |
|  | 4 | 5 (4.5, 5) | 4.5 (4.5, 4.5) | 3 (3, 3) |
|  | 5 (Least deprived) | 5 (5, 5) | 4.5 (4.5, 4.5) | 3 (3, 3) |

\*Limited to participants in work/education at baseline

## Discussion

In this study of over 20,000 adults from the Virus Watch cohort, the number of people who intended to accept, or had already accepted, a COVID-19 vaccine when offered increased from 90% in December 2020 to 97% in February 2021. Over four in five adults (86%) who were uncertain or intending to refuse a COVID-19 vaccine in December 2020 had changed their mind and planned on accepting, or had already accepted, a vaccine in February 2021. This shift was consistent across all ethnic groups measured and all levels of social deprivation. Despite this shift, some disparities in vaccine intention still exist. Young adults, and people from Black and White Other ethnic backgrounds, were more likely to intend to refuse or be unsure about taking a COVID-19 vaccine, compared to older adults and White British people.

Both concerns about COVID-19 vaccines and concerns about COVID-19 illness predicted intention to take a vaccine. Older adults were the least likely to have concerns about the vaccine and most likely to have concerns about COVID-19. White British people had fewer concerns about COVID-19 vaccines than people from most minority ethnic groups, but there were no differences between ethnic groups regarding concerns about contracting and/or becoming unwell with COVID-19 illness.

Our analysis of the Virus Watch cohort that examined how intention to take or refuse a COVID-19 vaccine has changed over time in England and Wales. It provides evidence that individuals are changing their minds and supports cross-sectional surveys that have found vaccination intention has increased over time.^21,22^ Previous studies have found age, ethnicity and social deprivation are independently associated with intention to take a COVID-19 vaccine.^9,10^ This study confirms that younger adults were far more likely to be unsure or intend to refuse a COVID-19 vaccine. This study finds that the association between ethnic group and vaccine intention has weakened. In February 2021, only people from Black ethnic backgrounds were more likely to refuse, and only people from White Other ethnic groups were more likely to be unsure about taking a COVID-19 vaccine compared to White British people. This is a substantial change from December 2021 and differs from existing evidence on ethnic disparities in COVID-19 vaccine intention.^9,10^ Similarly, the inverse relationship between deprivation and vaccine intention (where more deprived groups were associated with lower levels of intention to be vaccinated) has weakened, but not disappeared.

Beliefs and concerns about COVID-19 vaccines were the strongest predictor of intended vaccine uptake. This is in keeping with evidence from an earlier study conducted between September and October 2020 (prior to the UK COVID-19 vaccination programme commencing) which found that beliefs around the efficacy, development, risks and importance of COVID-19 vaccines strongly predicted intention to accept a vaccine.^15^ Our study also found beliefs and concerns around COVID-19 illness consistently predict vaccination intention. Our analysis was not able to consider structural reasons that determine vaccination intention beyond the role of missing work and education. Specifically, we did not measure conspiratorial beliefs and views of healthcare and medicine which have been previously found to predict vaccination intention to some degree.

Virus Watch is a national household community cohort study. Individuals in the study were geographically distributed across England and Wales and the cohort was diverse in terms of age, sex, ethnicity, and socio-economic composition. To our knowledge, this is the only cohort study of vaccination intention in England and Wales and has the largest number of participants from minority ethnic backgrounds. However, given participation in the Virus Watch study is voluntary and sampling non-random, the cohort is likely biased toward people concerned about COVID-19 and participants may not be representative of the national population. This may lead to an overestimate of public vaccination intention. This selection bias may be more apparent in groups hitherto associated with uncertainty or intended refusal of a COVID-19 vaccine, due to structural differences in access to and participation in healthcare and medical research.^23–25^

Nevertheless, the magnitude and consistency of the change in vaccine intention observed in this study is likely to outweigh possible bias. Despite the cohort size, samples were too small to disaggregate ethnic groups into more granular categories. Guided by our community advisory board, participants who expressed uncertainty were separated from participants intending to refuse COVID-19 vaccines in our regression analyses, as it was felt that these groups characterise different populations. Sample sizes precluded this separation in our analysis of temporal trends and analysis of psychological influences on vaccination intention. Another important limitation is that only households with a lead householder able to speak English were able to take part in the study and that all participants must have access to the internet. Guided by the advice from our community advisory board, the study surveys have since been translated into 9 languages to allow non-English speakers to participate.

The increase in intention to accept a COVID-19 vaccine when offered observed in this study is encouraging. It is possible that public health communications to promote vaccination uptake have contributed to this shift; it is also possible that growing numbers of people being vaccinated during the period of this study has contributed to the change in intention. It is particularly encouraging that the intention to accept a COVID-19 vaccine has increased consistently across ethnic and social groups. Our findings show the importance of making repeated offers of a COVID-19 vaccine because many people have changed their mind over the course of a few months. Our findings also suggest that communications focussing on COVID-19 vaccine safety and effectiveness may be more effective than those focussing on COVID-19 illness risk and perception.

Disparities in vaccination coverage should not be conflated with disparities in vaccination intention. If differences in vaccination coverage persist between ethnic groups and areas of social deprivation, our findings suggest they may not be fully explained by differences in vaccination intention. Instead they may plausibly be an example of the inverse care law.^14^ Ensuring easy and local vaccine access through community participation, for example using local places of worship as vaccination centres, would help overcome some of the structural barriers to healthcare and vaccine access.^26^

High levels of uncertainty or intended refusal of a COVID-19 vaccine in young people, driven by low-risk perception of COVID-19 illness and greater concern about vaccine safety, may limit the ability of the UK’s vaccination programme to suppress community transmission. It is possible that the increasing availability of vaccines to younger age groups will lead to increased vaccination intention in young adults. As the UK’s COVID-19 vaccination programme advances to offering vaccines to younger adults, efforts to increase vaccination intention should be responsive to age group-specific concerns and perspectives.

## Data Availability

We aim to share aggregate data from this project on our website and via a "Findings so far" section on our website - https://ucl-virus-watch.net/. We will also be sharing individual record level data with personal identifiers removed on a research data sharing service such as the Office of National Statistics Secure Research Service. In sharing the data we will work within the principles set out in the UKRI Guidance on best practice in the management of research data. Access to use of the data whilst research is being conducted will be managed by the Chief Investigators (ACH and RWA) in accordance with the principles set out in the UKRI guidance on best practice in the management of research data. It is the intention that the data arising from this research will initially be collected, cleaned and validated by the UCL research team and once this has been completed will be shared for wider use. We aim to make subsets of the data more rapidly available both on our study website and via the public facing dashboard during the ongoing phase of data collection. In line with Principle 5 of the UKRI guidance on best practice in the management of research data, we plan to release data in batches as they become available or as updated results are published. Individual record data linked using NHS Digital will not be shared, only aggregated results. HES and mortality data may be obtained from a third party and are not publicly available. These data are owned by a third party and can be accessed by researchers applying to the Health and Social Care Information Centre for England. We will put analysis code on publicly available repositories to enable their reuse.

