## Supplementary Appendices for "Trends, patterns and psychological influences on COVID-19 vaccination intention: findings from a large prospective community cohort study in England and Wales (Virus Watch)"

### SUPPLEMENTARY MATERIALS

#### Appendix I - Factors associated with vaccination intention

##### IA. Construction of explanatory variables

- Age (on study entry) - grouped into: 16-24, 25-34, 35-44, 45-54, 55-64, 65-74, and 75+ years.
- Sex (at birth) - male or female; other options were excluded due to small numbers.
- Region of residence (England and Wales) - East Midlands, East of England, London, North East, North West, South East, South West, West Midlands, Yorkshire and The Humber, and Wales; assigned based on home postcode at baseline.
- Indices of Multiple Deprivation - composite metric of area-level deprivation; assigned based on home postcode, and grouped by quintile.
- Ethnicity - using ONS categories and grouped as follows: Black, Mixed, South Asian, Other Asian, White British, White Irish, White Other, and Other Ethnicity.
- UK Joint Committee on Vaccination and Immunisation's (JCVI's) COVID-19 vaccine priority groups:
    1. *Care home residents and staff - not defined separately due to inadequate numbers of staff and lack of residents.*
    2. Age 80 years and over; or health or care worker (including care home staff, n=75).
    3. Age 75 to 79 years
    4. Age 70 to 74 years or clinically extremely vulnerable - the latter was defined using a proxy measure (response '*Yes*' to baseline survey question '*Have you received a letter from the NHS, saying that "the NHS has identified you as someone at risk of severe illness if you catch coronavirus, because you have an underlying disease or health condition that means if you catch the virus, you are more likely to be admitted to hospital than others"?*')
       This does not include anyone newly defined as extremely clinically vulnerable due to expansion of the criteria in February 2021.
    5. Age 65 to 69 years.
    6. Age 16 to 64 years with a priority health condition* (see below)
    7. Age 60 to 64 years.
    8. Age 55 to 59 years.
    9. Age 50 to 54 years.

* Priority health condition criteria were adapted from the Green Book Chapter 14a version 4 (31 December 2020) to align with yes/no questions on presence of pre-existing health conditions from the Virus Watch baseline survey.
The included conditions were determined as follows:

- Chronic respiratory disease: included asthma (severe asthma could not be distinguished, therefore all asthma was included); COPD; emphysema; chronic bronchitis; or cystic fibrosis. Other specified lung conditions could not be ascertained.

- Chronic heart disease and vascular disease: included angina; coronary heart disease; myocardial infarction; congestive heart failure; or hypertension (hypertension with cardiac complications could not be distinguished so all hypertension was included).
- Chronic kidney disease: included chronic kidney disease (stages could not be distinguished so all CKD was included).
- Chronic liver disease: those responding ‘Yes’ to ‘*any kind of liver condition*’ were included.
- Chronic neurological disease: those responding ‘Yes’ to *‘conditions affecting brain and nerves such as Parkinson’s, motor neurone disease, multiple sclerosis, learning disability, cerebral palsy’* were included; and separately, multiple sclerosis, epilepsy, and stroke were included.
- Diabetes mellitus: those responding ‘Yes’ to ‘*insulin treated diabetes*’ or ‘*other diabetes*’ were included.
- Immunosuppression: included immunosuppressive medication; malignancy; and HIV.
- Asplenia or dysfunction of the spleen:those responding ‘Yes’ to ‘*problems with your spleen or... spleen removed*’ were included; and separately, sickle cell disease was included.
- Morbid obesity: those with BMI ≥40 kg/m² were included; this was derived from height (cm) and weight (kg) reported at baseline. Implausible values (12>BMI>50), assessed as likely to be due to misinterpretation of units, were excluded.

Conditions or criteria that could not be ascertained or included in their entirety:

- Severe mental illness: not included as could not be distinguished from common mental illness. All mental illness was not included due to its high prevalence.
- Specific genetic disorders, e.g. those causing immunosuppression or splenic disorders, could not be ascertained. They may be included as part of other conditions/criteria but some may be missed.
- Specificer haematological disorders beyond those listed above could not be ascertained. They may be included as part of other conditions/criteria but some may be missed.
- Adult carers: not included as this could not be ascertained.
- Younger adults in long-stay nursing and residential care settings: not included as could not be ascertained, though these individuals are very unlikely to be in the Virus Watch cohort.

IB. Description of participant responses to ‘Would you accept a COVID-19 vaccine if offered?’ in December 2020 and February 2021 by socio-demographic factor

| | December | | | | February | | | | |
|---|---|---|---|---|---|---|---|---|---|
| | Total | Yes | Unsure | No | Total | Already had | Yes | Unsure | No |
| | N=20,792 | N=18,517 | N=1,813 | N=462 | N=20,284 | N=7,778 | N=12,039 | N=284 | N=183 |
| **Sex** (at birth) | | | | | | | | | |
| Male | 7,949 (100%) | 7,245 (91%) | 556 (7%) | 148 (2%) | 8,210 (100%) | 3,291 (40%) | 4,753 (58%) | 106 (1%) | 60 (1%) |
| Female | 10,399 (100%) | 9,162 (88%) | 1,020 (10%) | 217 (2%) | 9,092 (100%) | 3,554 (39%) | 5,340 (59%) | 116 (1%) | 82 (1%) |
| Other or missing | 2,444 (100%) | 2,110 (86%) | 237 (10%) | 97 (4%) | 2,982 (100%) | 933 (31%) | 1,946 (65%) | 62 (2%) | 41 (1%) |
| **Age** (years) | | | | | | | | | |
| 16-24 | 970 (100%) | 743 (77%) | 160 (16%) | 67 (7%) | 1,163 (100%) | 55 (5%) | 997 (86%) | 90 (8%) | 21 (2%) |
| 25-34 | 1,251 (100%) | 1,020 (82%) | 183 (15%) | 48 (4%) | 1,277 (100%) | 158 (12%) | 1,036 (81%) | 48 (4%) | 35 (3%) |
| 35-44 | 1,909 (100%) | 1,554 (81%) | 286 (15%) | 69 (4%) | 1,881 (100%) | 252 (13%) | 1,534 (82%) | 49 (3%) | 46 (2%) |
| 45-54 | 2,885 (100%) | 2,448 (85%) | 349 (12%) | 88 (3%) | 2,798 (100%) | 439 (16%) | 2,263 (81%) | 52 (2%) | 44 (2%) |
| 55-64 | 5,239 (100%) | 4,721 (90%) | 428 (8%) | 90 (2%) | 5,057 (100%) | 1,213 (24%) | 3,790 (75%) | 30 (1%) | 24 (0%) |
| 65-74 | 6,660 (100%) | 6,268 (94%) | 316 (5%) | 76 (1%) | - (100%) | - (69%) | - (30%) | - (0%) | - (0%) |
| 75+ | 1,878 (100%) | 1,763 (94%) | 91 (5%) | 24 (1%) | - (100%) | - (71%) | - (28%) | - (0%) | - (0%) |
| **Place of birth** | | | | | | | | | |
| Born in UK | 16,720 (100%) | 15,052 (90%) | 1,368 (8%) | 300 (2%) | 15,552 (100%) | 6,341 (41%) | 8,929 (57%) | 172 (1%) | 110 (1%) |
| Born abroad | 1,658 (100%) | 1,383 (83%) | 209 (13%) | 66 (4%) | 1,774 (100%) | 519 (29%) | 1,173 (66%) | 50 (3%) | 32 (2%) |
| Missing | 2,414 (100%) | 2,082 (86%) | 236 (10%) | 96 (4%) | 2,958 (100%) | 918 (31%) | 1,937 (65%) | 62 (2%) | 41 (1%) |
| **Ethnicity** | | | | | | | | | |
| White British | 16,506 (100%) | 14,915 (90%) | 1,305 (8%) | 286 (2%) | 15,373 (100%) | 6,340 (41%) | 8,770 (57%) | 159 (1%) | 104 (1%) |
| White Irish | - (100%) | - (88%) | - (10%) | - (1%) | - (100%) | - (40%) | - (57%) | - (1%) | - (2%) |

| | | | | | | | | | |
|---|---|---|---|---|---|---|---|---|---|
| White Other | 835 (100%) | 680 (81%) | 117 (14%) | 38 (5%) | 813 (100%) | 197 (24%) | 567 (70%) | 35 (4%) | 14 (2%) |
| South Asian | 300 (100%) | 242 (81%) | 43 (14%) | 15 (5%) | - (100%) | - (27%) | - (70%) | - (2%) | - (2%) |
| Other Asian | - (100%) | - (80%) | - (17%) | - (3%) | - (100%) | - (27%) | - (68%) | - (2%) | - (3%) |
| Black | - (100%) | - (60%) | - (31%) | - (9%) | - (100%) | - (20%) | - (71%) | - (5%) | - (5%) |
| Mixed | - (100%) | - (83%) | - (13%) | - (4%) | - (100%) | - (19%) | - (76%) | - (3%) | - (2%) |
| Other ethnicity | - (100%) | - (75%) | - (18%) | - (7%) | - (100%) | - (35%) | - (61%) | - (1%) | - (3%) |
| Prefer not to disclose or missing | 2,450 (100%) | 2,110 (86%) | 243 (10%) | 97 (4%) | 2,991 (100%) | 928 (31%) | 1,956 (65%) | 65 (2%) | 42 (1%) |
| **Region** | | | | | | | | | |
| East Midlands | 1,851 (100%) | 1,657 (90%) | 149 (8%) | 45 (2%) | 1,698 (100%) | 642 (38%) | 1,024 (60%) | 18 (1%) | 14 (1%) |
| East of England | 3,986 (100%) | 3,585 (90%) | 322 (8%) | 79 (2%) | 4,358 (100%) | 1,724 (40%) | 2,550 (59%) | 45 (1%) | 39 (1%) |
| London | 2,351 (100%) | 1,974 (84%) | 301 (13%) | 76 (3%) | 2,250 (100%) | 722 (32%) | 1,437 (64%) | 61 (3%) | 30 (1%) |
| North East | 1,054 (100%) | 945 (90%) | 84 (8%) | 25 (2%) | 922 (100%) | 318 (34%) | 581 (63%) | 11 (1%) | 12 (1%) |
| North West | 2,313 (100%) | 2,035 (88%) | 217 (9%) | 61 (3%) | 2,030 (100%) | 826 (41%) | 1,134 (56%) | 44 (2%) | 26 (1%) |
| South East | 3,464 (100%) | 3,145 (91%) | 277 (8%) | 42 (1%) | 3,637 (100%) | 1,448 (40%) | 2,124 (58%) | 37 (1%) | 28 (1%) |
| South West | 1,613 (100%) | 1,481 (92%) | 104 (6%) | 28 (2%) | - (100%) | - (41%) | - (58%) | - (1%) | - (0%) |
| West Midlands | 1,176 (100%) | 1,064 (90%) | 86 (7%) | 26 (2%) | - (100%) | - (42%) | - (55%) | - (2%) | - (0%) |
| Yorkshire and The Humber | 1,088 (100%) | 970 (89%) | 94 (9%) | 24 (2%) | - (100%) | - (40%) | - (58%) | - (2%) | - (0%) |
| Wales | 481 (100%) | 426 (89%) | 40 (8%) | 15 (3%) | - (100%) | - (34%) | - (65%) | - (1%) | - (0%) |
| Missing | 1,415 (100%) | 1,235 (87%) | 139 (10%) | 41 (3%) | 1,582 (100%) | 553 (35%) | 984 (62%) | 25 (2%) | 20 (1%) |
| **IMD** (quintiles) | | | | | | | | | |
| 1 (poorest) | 1,024 (100%) | 865 (84%) | 123 (12%) | 36 (4%) | - (100%) | - (32%) | - (65%) | - (2%) | - (1%) |
| 2 | 2,260 (100%) | 1,900 (84%) | 281 (12%) | 79 (3%) | 1,946 (100%) | 604 (31%) | 1,245 (64%) | 64 (3%) | 33 (2%) |
| 3 | 3,808 (100%) | 3,382 (89%) | 333 (9%) | 93 (2%) | 3,615 (100%) | 1,314 (36%) | 2,214 (61%) | 46 (1%) | 41 (1%) |
| 4 | 5,556 (100%) | 5,017 (90%) | 434 (8%) | 105 (2%) | 5,503 (100%) | 2,240 (41%) | 3,159 (57%) | 59 (1%) | 45 (1%) |
| 5 (richest) | 6,729 (100%) | 6,118 (91%) | 503 (7%) | 108 (2%) | 6,805 (100%) | 2,802 (41%) | 3,895 (57%) | 72 (1%) | 36 (1%) |

| | | | | | | | | | |
|---|---|---|---|---|---|---|---|---|---|
| Missing | 1,415 (100%) | 1,235 (87%) | 139 (10%) | 41 (3%) | - (100%) | - (35%) | - (62%) | - (2%) | - (1%) |
| **Health or care worker status** | 894 (100%) | 784 (88%) | 88 (10%) | 22 (2%) | - (100%) | - (63%) | - (36%) | - (0%) | - (1%) |
| **Priority health condition*** | 9,251 (100%) | 8,440 (91%) | 676 (7%) | 135 (1%) | 8,363 (100%) | 4,171 (50%) | 4,092 (49%) | 51 (1%) | 49 (1%) |
| **JCVI COVID-19 vaccine priority groups** | | | | | | | | | |
| Group 2<br>(health and care worker** or aged 80+ years) | 1,446 (100%) | 1,301 (90%) | 116 (8%) | 29 (2%) | - (100%) | - (67%) | - (32%) | - (0%) | - (1%) |
| Group 3<br>(aged 75-79 years) | 1,318 (100%) | 1,239 (94%) | 62 (5%) | 17 (1%) | - (100%) | - (71%) | - (29%) | - (0%) | - (0%) |
| Group 4<br>(clinically extremely vulnerable*** or aged 70-74 years) | 3,866 (100%) | 3,625 (94%) | 189 (5%) | 52 (1%) | - (100%) | - (72%) | - (28%) | - (0%) | - (0%) |
| Group 5<br>(aged 65-69 years) | 3,212 (100%) | 3,010 (94%) | 164 (5%) | 38 (1%) | - (100%) | - (64%) | - (35%) | - (0%) | - (0%) |
| Group 6<br>(higher risk health condition* and aged 16-64 years) | 3,472 (100%) | 3,011 (87%) | 391 (11%) | 70 (2%) | 3,191 (100%) | 553 (17%) | 2,558 (80%) | 43 (1%) | 37 (1%) |
| Group 7<br>(aged 60-64 years) | 1,515 (100%) | 1,379 (91%) | 107 (7%) | 29 (2%) | - (100%) | - (20%) | - (79%) | - (0%) | - (0%) |
| Group 8<br>(aged 55-59 years) | 1,230 (100%) | 1,084 (88%) | 120 (10%) | 26 (2%) | - (100%) | - (14%) | - (84%) | - (1%) | - (1%) |
| Group 9<br>(aged 50-54 years) | 941 (100%) | 822 (87%) | 94 (10%) | 25 (3%) | 956 (100%) | 100 (10%) | 818 (86%) | 22 (2%) | 16 (2%) |
| Not in any JCVI priority group | 3,792 (100%) | 3,046 (80%) | 570 (15%) | 176 (5%) | 4,019 (100%) | 258 (6%) | 3,485 (87%) | 181 (5%) | 95 (2%) |

**Due to low cell counts across certain categories of explanatory variables, the data with "-" have been suppressed to reduce the possibility of deductive disclosure.*

IC. December Survey - multivariable regression analysis results comparing demographic and social factors of those responding ‘No’ and those responding ‘Unsure’ with those responding ‘Yes’ when asked ‘Would you accept a COVID-19 vaccine if offered?’

| | 'No' | | | | | | | 'Unsure' | | | | | | |
|---|---|---|---|---|---|---|---|---|---|---|---|---|---|---|
| | Partially adjusted (age and gender) | | | **Fully adjusted multivariable model (n=16,617)** | | | | Partially adjusted (age, gender) | | | **Fully adjusted multivariable model (n=17,812)** | | | |
| | **RR** | **95% CI** | | **RR** | **95% CI** | | **p-value** | **RR** | **95% CI** | | **RR** | **95% CI** | | **p-value** |
| **Female** | 1.13 | 0.92 | 1.39 | 1.11 | 0.94 | 1.31 | 0.226 | 1.35 | 1.23 | 1.49 | 1.34 | 1.24 | 1.44 | 0.000 |
| **Age (years)** | | | | | | | | | | | | | | |
| **16-24** | 5.64 | 3.29 | 9.66 | 4.32 | 2.40 | 7.78 | <0.001 | 3.30 | 2.50 | 4.36 | 2.93 | 2.19 | 3.92 | <0.001 |
| **25-34** | 3.24 | 1.87 | 5.63 | 2.26 | 1.22 | 4.19 | 0.010 | 3.04 | 2.34 | 3.94 | 2.61 | 1.96 | 3.48 | <0.001 |
| **35-44** | 3.84 | 2.31 | 6.38 | 2.72 | 1.56 | 4.75 | <0.001 | 3.20 | 2.51 | 4.08 | 2.81 | 2.16 | 3.66 | <0.001 |
| **45-54** | 2.46 | 1.48 | 4.10 | 2.16 | 1.25 | 3.73 | 0.006 | 2.54 | 2.00 | 3.23 | 2.28 | 1.77 | 2.94 | <0.001 |
| **55-64** | 1.39 | 0.84 | 2.30 | 1.22 | 0.72 | 2.08 | 0.455 | 1.68 | 1.33 | 2.13 | 1.62 | 1.27 | 2.08 | <0.001 |
| **65-74** | 0.93 | 0.56 | 1.55 | 0.86 | 0.51 | 1.43 | 0.555 | 0.97 | 0.76 | 1.23 | 0.95 | 0.74 | 1.21 | 0.676 |
| **75+** | 1 | | | 1 | | | - | 1 | | | 1 | | | - |
| **Ethnicity** | | | | | | | | | | | | | | |
| **White British** | 1 | | | 1 | | | - | 1 | | | 1 | | | - |
| **White Irish** | 0.70 | 0.23 | 2.16 | 0.63 | 0.22 | 1.85 | 0.403 | 1.21 | 0.83 | 1.76 | 1.21 | 0.82 | 1.77 | 0.330 |
| **White Other** | 2.05 | 1.45 | 2.89 | 2.07 | 1.38 | 3.08 | <0.001 | 1.35 | 1.13 | 1.61 | 1.35 | 1.11 | 1.65 | <0.001 |
| **South Asian** | 2.21 | 1.32 | 3.70 | 2.52 | 1.26 | 5.06 | 0.009 | 1.47 | 1.10 | 1.96 | 1.58 | 1.07 | 2.33 | 0.021 |
| **Other Asian** | 1.31 | 0.51 | 3.39 | 0.96 | 0.45 | 2.03 | 0.912 | 1.52 | 1.03 | 2.25 | 1.59 | 1.04 | 2.42 | 0.033 |
| **Black** | 4.92 | 2.34 | 10.31 | 4.78 | 1.98 | 11.51 | <0.001 | 3.38 | 2.42 | 4.73 | 3.57 | 2.36 | 5.40 | <0.001 |
| **Mixed** | 1.66 | 0.83 | 3.30 | 1.35 | 0.72 | 2.53 | 0.355 | 1.14 | 0.78 | 1.66 | 1.10 | 0.78 | 1.57 | 0.581 |
| **Other ethnicity** | 3.99 | 1.79 | 8.93 | 2.80 | 1.16 | 6.74 | 0.022 | 2.17 | 1.37 | 3.44 | 1.89 | 1.09 | 3.26 | 0.023 |

| | | | | | | | | | | | | | | |
|---|---|---|---|---|---|---|---|---|---|---|---|---|---|---|
| **IMD quintiles** | | | | | | | | | | | | | | |
| **1 (poorest)** | 2.29 | 1.56 | 3.34 | 2.30 | 1.37 | 3.87 | 0.002 | 1.49 | 1.23 | 1.81 | 1.60 | 1.26 | 2.04 | <0.001 |
| **2** | 1.88 | 1.38 | 2.56 | 1.78 | 1.20 | 2.64 | 0.004 | 1.44 | 1.25 | 1.66 | 1.45 | 1.21 | 1.74 | <0.001 |
| **3** | 1.43 | 1.07 | 1.92 | 1.30 | 0.89 | 1.89 | 0.168 | 1.09 | 0.95 | 1.25 | 1.09 | 0.92 | 1.30 | 0.311 |
| **4** | 1.01 | 0.76 | 1.36 | 1.02 | 0.72 | 1.46 | 0.899 | 1.02 | 0.90 | 1.15 | 1.02 | 0.87 | 1.20 | 0.823 |
| **5 (richest)** | 1 | | | 1 | | | | 1 | | | 1 | | | |
| **Region** | | | | | | | | | | | | | | |
| **East Midlands** | 1.22 | 0.84 | 1.79 | | | | | 1.04 | 0.86 | 1.26 | | | | |
| **East of England** | 1 | | | | | | | 1 | | | | | | |
| **London** | 1.35 | 0.96 | 1.90 | | | | | 1.35 | 1.15 | 1.58 | | | | |
| **North East** | 1.04 | 0.63 | 1.72 | | | | | 1.03 | 0.81 | 1.30 | | | | |
| **North West** | 1.27 | 0.89 | 1.81 | | | | | 1.15 | 0.97 | 1.36 | | | | |
| **South East** | 0.57 | 0.38 | 0.85 | | | | | 0.97 | 0.83 | 1.14 | | | | |
| **South West** | 0.89 | 0.57 | 1.41 | | | | | 0.84 | 0.68 | 1.05 | | | | |
| **West Midlands** | 1.06 | 0.65 | 1.71 | | | | | 0.93 | 0.74 | 1.18 | | | | |
| **Yorkshire and The Humber** | 1.23 | 0.77 | 1.95 | | | | | 1.09 | 0.87 | 1.37 | | | | |
| **Wales** | 1.75 | 0.99 | 3.10 | | | | | 1.05 | 0.75 | 1.46 | | | | |
| **Health or care worker** | 0.98 | 0.63 | 1.52 | | | | | 0.86 | 0.70 | 1.06 | | | | |
| **Extremely clinically vulnerable*** | 1.19 | 0.79 | 1.80 | | | | | 0.90 | 0.73 | 1.11 | | | | |
| **Priority health condition*** | 0.78 | 0.62 | 0.97 | 0.77 | 0.62 | 0.96 | 0.019 | 0.96 | 0.87 | 1.06 | 0.94 | 0.85 | 1.03 | 0.178 |

**See Appendix IA*

ID. February Survey Multivariable - multivariable regression analysis results comparing demographic and social factors of participants responding ‘No’ and those responding ‘Unsure’ with those responding ‘Yes’ when asked ‘Would you accept a COVID-19 vaccine if offered?’

| | 'No' | | | | | | | 'Unsure' | | | | | | |
|---|---|---|---|---|---|---|---|---|---|---|---|---|---|---|
| | Partially adjusted (age and gender) | | | **Fully adjusted multivariable model** (n=16,927) | | | | Partially adjusted (age, gender) | | | **Fully adjusted multivariable model** (n=17,005) | | | |
| | **RR** | **95% CI** | | **RR** | **95% CI** | | p-value | **RR** | **95% CI** | | **RR** | **95% CI** | | p-value |
| **Female** | 1.09 | 0.78 | 1.52 | 1.24 | 0.95 | 1.60 | 0.108 | 0.92 | 0.71 | 1.20 | 0.92 | 0.72 | 1.18 | 0.511 |
| **Age** (years) | | | | | | | | | | | | | | |
| **16-24** | 7.44 | 2.10 | 26.29 | 5.30 | 1.39 | 20.20 | 0.014 | 27.16 | 9.90 | 74.55 | 20.21 | 7.19 | 56.78 | <0.001 |
| **25-34** | 12.55 | 3.83 | 41.17 | 8.58 | 2.35 | 31.39 | 0.001 | 13.18 | 4.70 | 36.95 | 7.90 | 2.71 | 23.04 | <0.001 |
| **35-44** | 11.64 | 3.60 | 37.59 | 7.47 | 2.08 | 26.77 | 0.002 | 9.45 | 3.39 | 26.39 | 5.62 | 1.97 | 16.06 | 0.001 |
| **45-54** | 6.54 | 2.01 | 21.31 | 4.71 | 1.31 | 16.91 | 0.018 | 6.73 | 2.41 | 18.77 | 4.69 | 1.65 | 13.34 | 0.004 |
| **55-64** | 2.28 | 0.68 | 7.63 | 1.80 | 0.49 | 6.59 | 0.374 | 2.10 | 0.73 | 6.04 | 1.76 | 0.61 | 5.10 | 0.299 |
| **65-74** | 0.58 | 0.15 | 2.26 | 0.47 | 0.11 | 2.02 | 0.311 | 0.64 | 0.20 | 2.03 | 0.54 | 0.17 | 1.77 | 0.310 |
| **75+** | 1 | | | 1 | | | | 1 | | | 1 | | | |
| **Ethnicity** | | | | | | | | | | | | | | |
| **White British** | 1 | | | 1 | | | | 1 | | | 1 | | | |
| **White Irish** | 2.31 | 0.88 | 6.07 | 1.79 | 0.61 | 5.25 | 0.290 | 1.46 | 0.47 | 4.54 | 1.40 | 0.48 | 4.04 | 0.538 |
| **White Other** | 1.34 | 0.77 | 2.34 | 1.30 | 0.71 | 2.41 | 0.394 | 2.56 | 1.76 | 3.71 | 2.58 | 1.67 | 3.98 | <0.001 |
| **South Asian** | 1.37 | 0.64 | 2.91 | 1.12 | 0.48 | 2.61 | 0.787 | 1.16 | 0.59 | 2.25 | 1.01 | 0.43 | 2.36 | 0.985 |
| **Other Asian** | 2.85 | 1.07 | 7.59 | 2.16 | 0.70 | 6.67 | 0.180 | 1.40 | 0.46 | 4.26 | 1.28 | 0.44 | 3.72 | 0.654 |
| **Black** | 3.78 | 1.26 | 11.33 | 5.45 | 1.46 | 20.32 | 0.012 | 2.44 | 0.77 | 7.77 | 2.97 | 0.75 | 11.67 | 0.119 |
| **Mixed** | 1.21 | 0.40 | 3.69 | 0.84 | 0.30 | 2.35 | 0.736 | 1.52 | 0.68 | 3.41 | 1.63 | 0.70 | 3.81 | 0.261 |
| Other ethnicity | 2.86 | 0.70 | 11.62 | 2.41 | 0.70 | 8.33 | 0.165 | 0.99 | 0.15 | 6.34 | 0.78 | 0.14 | 4.39 | 0.782 |
| **IMD quintiles** | | | | | | | | | | | | | | |

| | | | | | | | | | | | | | | |
|---|---|---|---|---|---|---|---|---|---|---|---|---|---|---|
| **1 (poorest)** | 1.64 | 0.76 | 3.54 | 2.11 | 0.86 | 5.20 | 0.105 | 1.89 | 1.12 | 3.19 | 2.02 | 1.04 | 3.95 | 0.039 |
| **2** | 2.22 | 1.35 | 3.65 | 2.62 | 1.38 | 4.97 | 0.003 | 2.38 | 1.64 | 3.46 | 2.38 | 1.50 | 3.77 | <0.001 |
| **3** | 1.77 | 1.10 | 2.84 | 2.03 | 1.10 | 3.77 | 0.024 | 0.99 | 0.66 | 1.49 | 0.99 | 0.61 | 1.58 | 0.953 |
| **4** | 1.38 | 0.86 | 2.22 | 1.39 | 0.76 | 2.55 | 0.284 | 1.13 | 0.78 | 1.62 | 1.08 | 0.70 | 1.67 | 0.735 |
| **5 (richest)** | 1 | | | 1 | | | | 1 | | | 1 | | | |
| **Region** | | | | | | | | | | | | | | |
| **East Midlands** | 0.83 | 0.42 | 1.66 | | | | | 0.92 | 0.51 | 1.63 | | | | |
| **East of England** | 1 | | | | | | | 1 | | | | | | |
| **London** | 1.05 | 0.62 | 1.77 | | | | | 1.61 | 1.06 | 2.46 | | | | |
| **North East** | 1.67 | 0.85 | 3.28 | | | | | 1.12 | 0.54 | 2.30 | | | | |
| **North West** | 1.54 | 0.91 | 2.61 | | | | | 1.81 | 1.16 | 2.80 | | | | |
| **South East** | 0.94 | 0.56 | 1.60 | | | | | 0.89 | 0.56 | 1.41 | | | | |
| **South West** | 0.52 | 0.20 | 1.34 | | | | | 0.57 | 0.26 | 1.26 | | | | |
| **West Midlands** | 0.57 | 0.20 | 1.61 | | | | | 1.63 | 0.92 | 2.89 | | | | |
| **Yorkshire and The Humber** | 0.66 | 0.23 | 1.86 | | | | | 1.63 | 0.88 | 3.00 | | | | |
| **Wales** | 0.37 | 0.05 | 2.75 | | | | | 0.49 | 0.12 | 2.00 | | | | |
| **Health or care worker** | 0.40 | 0.15 | 1.07 | | | | | 0.16 | 0.04 | 0.63 | | | | |
| **Extremely clinically vulnerable*** | 0.88 | 0.39 | 1.99 | | | | | 0.41 | 0.15 | 1.10 | | | | |
| **Priority health condition*** | 0.95 | 0.67 | 1.35 | 0.89 | 0.65 | 1.23 | 0.485 | 0.60 | 0.43 | 0.82 | 0.62 | 0.46 | 0.85 | 0.003 |

**See Appendix IA*

#### Appendix II - Psychological influences on COVID-19 vaccination intention

[Participant Name]: We would like to know your views about COVID-19 and about receiving a vaccine for yourself, if you were offered. Please indicate your agreement with each of the following statements:

**Item scoring:**

| 1 | 2 | 3 | 4 | 5 |
|---|---|---|---|---|
| Strongly Disagree | Disagree | Neither Agree nor Disagree | Agree | Strongly Agree |

**Items** (*note: * =reverse scored for analyses)*

1) I am worried that if I caught COVID-19 I might pass it on to others
2) COVID-19 is very serious if you catch it
3) *I do not think that I am at risk of COVID-19
4) *I do not think that I am at high risk of complications of COVID-19
5) *I am worried about having to take time off work/education because of COVID-19
6) COVID-19 vaccine would be safe for me
7) COVID-19 vaccine would be effective in preventing me from getting COVID- 19
8) *Natural infection provides me with stronger immunity
9) *I do not trust vaccines
10) *I am too busy/have too little time to get vaccinated
11) *I am concerned that the COVID-19 vaccine will not have been tested enough
12) *I am concerned that the vaccine will make you feel as ill as COVID-19 does
13) *I am concerned about rare but serious side effects of the COVID-19 vaccine

#### Appendix III - FACTOR ANALYSIS

We performed exploratory factor analysis (EFA) using the principal axis factoring method to account for skewed item distributions, with Promax rotation to account for plausible correlation in underlying factors.[13] Factors were extracted based on examination of scree plots and Kaiser's criterion. We then assessed absolute and relative model fit and compared to a univariate model using confirmatory factor analysis (CFA) with the Satorra-Bentler adjustment. We then computed participants' median response across items in each identified factor. Median factor scores were entered as exposures for univariable Poisson models for vaccination intention, as described above; we did not conduct multivariable analysis due to putative mediation effects.

Kaiser-Meyer-Olkin values (respectively 0.81 and 0.82) and Bartlett's tests ($p$<.001) suggested that both the EFA and CFA datasets were appropriate for factor analysis.

**Table 1. Exploratory Factor Analysis Results (n=10088)**
*Results in parentheses are prior to removal of Item 5*

| | $h^2$ | Factor Loading | Cronbach's α | Eigenvalue | Variance Explained |
|---|---|---|---|---|---|
| Factor 1 | - | - | 0.80 (0.80) | 3.19 (3.19) | 0.72 (0.72)§ |
| Item 6 | 0.45 (0.45) | 0.64 (0.63) | | | |
| Item 7 | 0.34 (0.34) | 0.54 (0.53) | | | |
| Item 8 | 0.10 (0.09) | 0.22 (0.21) | | | |
| Item 9 | 0.37 (0.37) | 0.58 (0.58) | | | |
| Item 10 | 0.22 (0.22) | 0.40 (0.40) | | | |
| Item 11 | 0.59 (0.59) | 0.77 (0.77) | | | |
| Item 12 | 0.55 (0.55) | 0.75 (0.76) | | | |
| Item 13 | 0.51 (0.52) | 0.74 (0.75) | | | |
| Factor 2 | - | - | 0.67 (0.67) | 1.73 (1.80) | 0.39 (0.40)§ |
| Item 1 | 0.23 (0.24) | 0.44 (0.46) | | | |
| Item 2 | 0.37 (0.37) | 0.58 (0.59) | | | |
| Item 3 | 0.39 (0.39) | 0.62 (0.63) | | | |
| Item 4 | 0.38 (0.38) | 0.63 (0.63) | | | |
| Total Scale | | | 0.78 (0.75) | | |

**Note:** $h^2$= communalities. Factor loadings >0.20 reported.
§Due to the correlation between factors ($r$=0.27) arising from the oblique rotation, explained variance for the two factors is non-independent and thus appears to exceed 100%.

**Table 2. Confirmatory Factor Analysis Results (*n*=10089)**

| | | | | | | | Standardised Factor Loadings | | | |
|---|---|---|---|---|---|---|---|---|---|---|
| | $X^2$ (df) | RMSEA (90% CI) | CFI | TLI | AIC | BIC | Univariate (all items) | Univariate (excl. Item 5) | EFA Factor 1 (EFA Loading) | EFA Factor 2 (EFA Loading) |
| Two-Factor EFA Model | 4858.75 (53) | 0.09 | 0.82 | 0.77 | 284500.12 | 284767.23 | | | | |
| Univariate model (all items) | 9392.59 (65) | 0.12 | 0.66 | 0.59 | 324594.66 | 324876.21 | | | | |
| Univariate model (excl. Item 5) | 8851.64 (54) | 0.13 | 0.66 | 0.59 | 290059.68 | 290319.57 | | | | |
| Item 1 | | | | | | | 0.23 | 0.24 | | 0.40 (0.44) |
| Item 2 | | | | | | | 0.26 | 0.26 | | 0.54 (0.58) |
| Item 3 | | | | | | | 0.21 | 0.21 | | 0.73 (0.62) |
| Item 4 | | | | | | | 0.17 | 0.17 | | 0.71 (0.63) |
| Item 5 | | | | | | | 0.12 | n/a | n/a | n/a |
| Item 6 | | | | | | | 0.63 | 0.63 | 0.61 (0.64) | |
| Item 7 | | | | | | | 0.55 | 0.55 | 0.53 (0.54) | |
| Item 8 | | | | | | | 0.29 | 0.29 | 0.28 (0.22) | |
| Item 9 | | | | | | | 0.59 | 0.59 | 0.59 (0.58) | |
| Item 10 | | | | | | | 0.45 | 0.45 | 0.44 (0.40) | |
| Item 11 | | | | | | | 0.79 | 0.79 | 0.80 (0.77) | |
| Item 12 | | | | | | | 0.75 | 0.75 | 0.77 (0.75) | |
| Item 13 | | | | | | | 0.72 | 0.71 | 0.74 (0.74) | |

**Note:** All factor loadings significant p<0.0001; EFA= exploratory factor analysis; RMSEA = Root Mean Square Error of Approximation (close fit: <.05, adequate fit <0.8)[1]; CFI = Comparative Fit Index (close fit: 0.9)[2]; TLI = Taylor-Lewis Index (close fit: >0.9)[3]; AIC = Akaike's Information Criterion (lower values indicating better fit)[4]; BIC = Bayesian Information Criterion (lower values indicating better fit)[5]
